# Decoding the regulatory genetic architecture of endometriosis using AlphaGenome

**DOI:** 10.64898/2026.06.27.26356730

**Authors:** Sanu Bifal Maji, Apostol Apostolov, Alberto Sola-Leyva, Lucia Blanco-Rodriguez, Amruta D. S. Pathare, Andres Salumets

**Author notes:** Corresponding author – Andres Salumets. Co-last authors (Amruta D. S. Pathare, Andres Salumets).

## Abstract

**Background:** Endometriosis is a complex, estrogen-dependent disease with a strong genetic component. Although genome-wide association studies (GWAS) have identified multiple susceptibility loci, most associated variants reside in noncoding regions, limiting biological interpretation and causal gene identification. Moreover, GWAS gene prioritization is limited by incomplete tissue-specific annotation coverage (e.g., GTEx, ENCODE, fine-mapping, Mendelian randomization, and network-based methods). We therefore applied the AlphaGenome artificial intelligence framework to prioritize endometriosis-associated variants based on predicted uterus-specific regulatory effects.

**Methods:** We analysed the top 10,000 endometriosis-associated single-nucleotide polymorphisms (SNPs) identified by previously published GWAS by Rahmioglu *et al*, using AlphaGenome across multiple genomic output types. Uterus-specific predictions with high-confidence effects (|quantile score| ≥ 0.90) were grouped into major regulatory modalities. AlphaGenome-prioritized SNPs within ±500 kb of known GWAS loci were classified into tiers based on the number of supported regulatory modalities, with broader support indicating stronger multilayer regulatory evidence. Effect allele frequency, linkage disequilibrium (LD), and overlap with previously published endometriosis-associated variants were also assessed.

**Results:** AlphaGenome generated uterus-specific, 147,033 high-confidence signals across 10,000 endometriosis-associated variants, spanning six regulatory modalities including gene expression, promoter activity, chromatin accessibility, transcription factor binding, histone modification, and RNA splicing. Within the 42 established endometriosis GWAS loci, AlphaGenome identified 42 alternative sub-threshold SNPs with stronger predicted uterus-specific regulatory effects than the published GWAS lead variants. Nineteen AlphaGenome-prioritized SNPs were classified as tier 1, showing support across all six regulatory modalities, compared with five GWAS lead SNPs. Linkage disequilibrium analysis identified eight tier 1 SNPs with weak-to-low LD (r² < 0.5) relative to the corresponding GWAS lead variants, regulating majority of genes involved in estrogen-driven proliferation and inflammatory signalling, highlighting their potential relevance to endometriosis pathogenesis. Additionally, we identified 167 genome-wide significant SNPs outside 42 published GWAS lead SNP loci including six tier 1 SNPs (rs1482061, rs7772579, rs6557140, rs2982571, rs12631337 and rs79626929), encompassing genes nearby *ESR1/*6q25.1, substantiating biological relevance for endometriosis pathogenesis.

**Conclusions:** AlphaGenome-based regulatory prioritization refined endometriosis-associated genome-wide association study loci by identifying variants with stronger predicted uterus-specific functional relevance. These findings provide a regulatory framework for prioritizing candidate variants and genes for downstream functional validation in endometriosis.

## Background

Endometriosis is a chronic inflammatory disorder affecting approximately 5-10% of women of reproductive age and is characterized by the presence of endometrial-like tissue outside the uterus, most commonly on pelvic organs [1]. The disease is associated with debilitating pelvic pain, infertility, and reduced quality of life, and diagnosis is often substantially delayed because definitive confirmation typically requires surgical visualization of lesions [2,3]. Current treatment options remain limited, relying mainly on hormonal suppression or invasive approaches to surgically remove the lesions, both of which have limitations and important impact on women health. Biologically and clinically, endometriosis is considered a heterogeneous condition characterized by variability in lesion types, disease stage, infertility, and pain manifestations, suggesting that multiple pathogenic mechanisms contribute to disease susceptibility and presentation.

Genetic factors make a major contribution to endometriosis risk, with heritability estimated at approximately 50%, with substantial proportion attributed to common genetic variation [4,5]. Recent large-scale genome-wide association studies (GWAS) have expanded understanding of the genetic architecture of endometriosis [4,6–13]. In particular, a meta-analysis including 60,674 cases and 701,926 controls identified 42 genome-wide significant endometriosis-related loci [3]. Fine-mapping of these loci further resolved six high-confidence candidate causal variants, including variants at or near *SYNE1, HOXA10, HOXC10, LINC00629, ESR1*, and *LNC-LBCS* genes, all located in non-coding regions [3]. More recently, a large multi-ancestry GWAS and integrated multi-omics analysis identified 80 genomic regions associated with endometriosis risk, including 37 newly reported loci [14]. Multi-omics integrative analyses in several tissues have further linked endometriosis genetic risk to pathways involved in cell differentiation, immune and hormonal regulation, tissue remodeling, and inflammation [3,14]. Consistent with these findings, many implicated loci map near genes involved in hormone signaling, uterine development, immune regulation, adhesion, and angiogenesis [1,7,9,10,12,13,15–25], which are vital biological processes related to the pathogenesis of endometriosis. However, despite these advances, the regulatory consequences of most endometriosis-associated variants remain incompletely understood, particularly because the majority reside in non-coding regions, making gene prioritization and mechanistic interpretation from GWAS signals alone inherently challenging. As a result, current GWAS-based approaches largely resolve association signals at the locus level rather than identifying causal genes or regulatory mechanisms, particularly in hormonally regulated tissues such as the uterus, where long-range and context-dependent gene regulation is prominent. To address this limitation, there is a need for approaches that can systematically link non-coding genetic variation to downstream regulatory effects across disease-relevant tissues and molecular layers.

In this context, recent advances in deep learning models such as AlphaGenome represent a major step forward in sequence-to-function modeling by enabling simultaneous prediction of thousands of functional genomic output tracks at base-pair resolution across multiple regulatory modalities, including gene expression, chromatin accessibility, transcription factor (TF) binding, histone modifications, promoter activity, three-dimensional chromatin organization and RNA splicing [26]. Unlike earlier models like SpliceAI [27], BPNet [28], ProCapNet [29], among others, which were designed for more specific regulatory tasks or were constrained by trade-offs between input sequence length and output resolution, AlphaGenome integrates long-range genomic context of up to 1 Mb with high-resolution output, enabling the modelling of both local and distal regulatory effects within a unified framework [26]. The model has demonstrated state-of-the-art or near state-of-the-art performance across multiple benchmarking tasks and has shown strong ability to predict the molecular consequences of non-coding variants across diverse regulatory layers [26]. Notably, AlphaGenome has also been shown to recapitulate known disease-associated regulatory mechanisms, including non-coding variant effects near the *TAL1* oncogene [30], where it successfully captured coordinated changes in TF binding, chromatin accessibility, and gene expression associated with oncogenic activation. Such performance highlights its value as a framework for linking non-coding genetic variation to downstream molecular consequences across multiple regulatory modalities [26]. Given that the majority of disease-associated GWAS variants as in endometriosis, lie in non-coding regions [3,5], tools such as AlphaGenome may provide an important opportunity to bridge the gap between genetic association and biological mechanism. This is particularly important for loci where fine-mapping, expression and methylation quantitative trait locus, or sub-phenotype analyses suggest functional relevance, yet the precise molecular direction and breadth of regulatory perturbation remain unresolved.

In this study, we integrated endometriosis GWAS risk loci [3,14] with uterus-specific AlphaGenome predictions to investigate the regulatory architecture of disease-associated variants across multiple molecular layers including transcription, promoter, chromatin accessibility, TF binding, histone modification, three-dimensional chromatin organization and RNA splicing. The variants based on this multimodal approach were scored to prioritize loci which could predict regulatory interpretation of endometriosis. By combining large-scale genetic association data with deep learning-based regulatory prediction, this work seeks to refine the functional interpretation of endometriosis risk loci and provide new insight into the tissue-relevant regulatory mechanisms underlying disease susceptibility and symptom heterogeneity.

## Methods

### Data source and variant processing

A total of 10,000 SNPs (*p* < 3.36 × 10 □^5^) were selected from the endometriosis GWAS meta-analysis reported in study by Rahmioglu *et al*, [3] (Additional file 2: Table S1). The initial variant information extracted from the GWAS source included chromosome, genomic position in hg19, and the corresponding effect and non-effect alleles. To ensure compatibility with AlphaGenome, which requires variant coordinates in the GRCh38/hg38 reference genome, all SNPs were converted from hg19 to hg38 using the Ensembl REST API. Variants were mapped individually, and only successfully converted variants were retained for downstream analysis. RsIDs were then annotated using Ensembl variation records based on hg38 genomic coordinates (Additional file 2: Table S2). For AlphaGenome prediction, variants were encoded such that the alternate allele corresponded to the GWAS effect allele, and the reference allele corresponded to the GWAS non-effect allele, after checking compatibility with the hg38 genomic position. The processed variants were formatted into a tab-separated input file compatible with AlphaGenome, including variant ID, chromosome in chr format, hg38 genomic position, reference/non-effect allele, and alternate/effect allele (Fig. 1, Additional file 1 and Additional file 2: Table S2). Results were further cross-referenced with a recent multi-ancestry endometriosis GWAS dataset [14] to investigate the regulatory modalities associated with these genetic signals.

**Figure 1.**
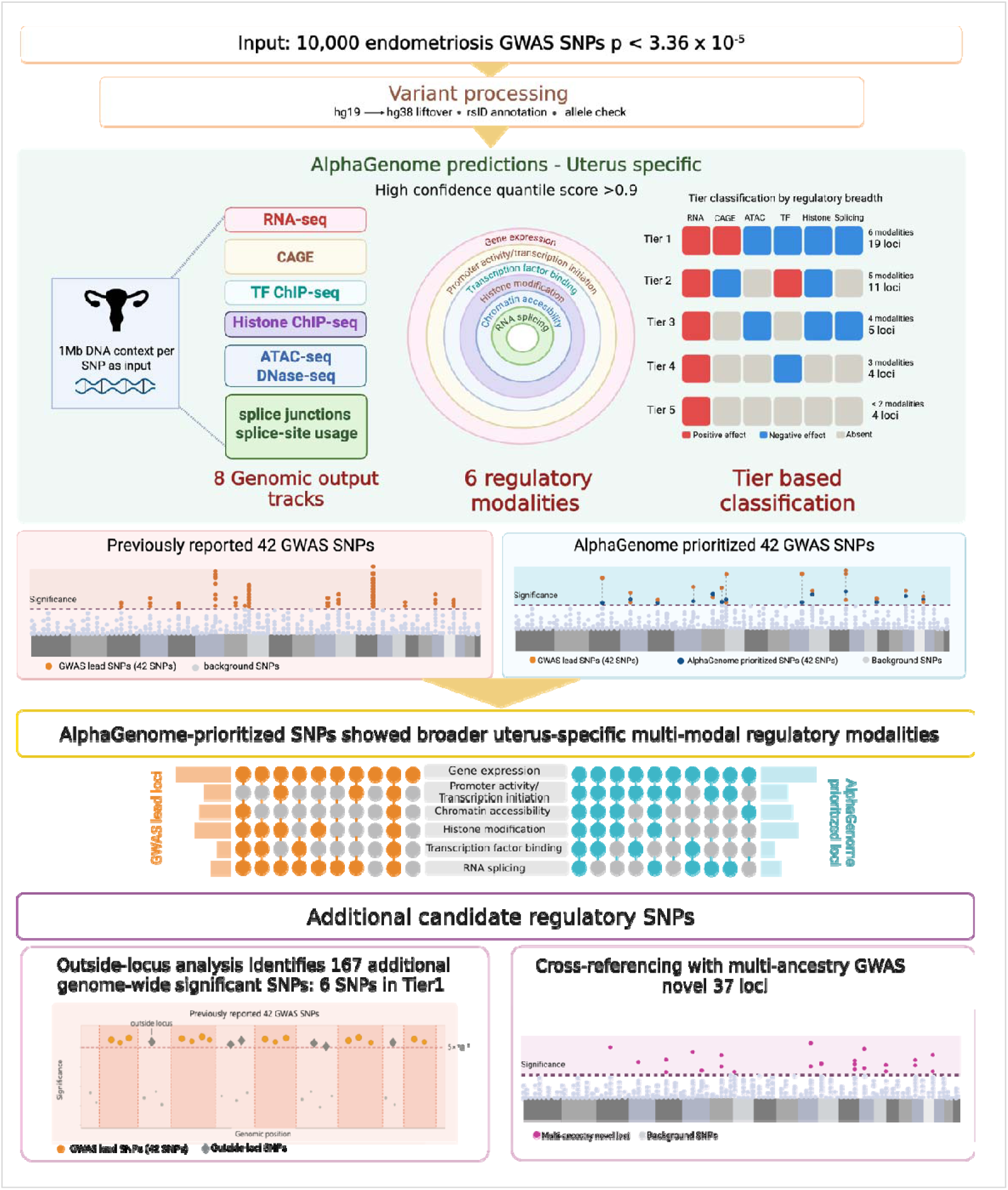
**AlphaGenome workflow for prioritizing endometriosis-associated SNPs**. A total of 10,000 endometriosis-associated GWAS SNPs (*p* < 3.36×10 □ □) were converted from hg19 to hg38 and analyzed using AlphaGenome with a 1 Mb sequence context across 11 regulatory genomic output types. Predictions were restricted to uterus-specific biosamples, and high-confidence signals (|quantile score| ≥ 0.90). Genomic output types were grouped into six regulatory modalities like gene expression, promoter activity, chromatin accessibility, transcription factor binding, histone modification, and RNA splicing. Each SNP was assigned a tier based on regulatory modalities (Tier 1 = all six modalities; Tier 5 = ≤2 modalities). Tiered SNPs were evaluated across three groups: published GWAS lead SNPs at 42 genome-wide significant loci, AlphaGenome-prioritized alternative SNPs within ±500 kb of these loci, and outside-locus SNPs. Results were further cross-referenced with a recent multi-ancestry endometriosis GWAS [14] to validate novel regulatory signals.

### AlphaGenome-based variant effect prediction

Variant effect prediction was performed using the AlphaGenome Python SDK version 0.6.1 within a Conda-based computational environment. The model was configured for *Homo sapiens* using a 1 Mb sequence context window, allowing both proximal and distal regulatory elements surrounding each variant to be incorporated into the prediction [26]. For each SNP, a genomic interval centered on the variant position was extracted and used as input to the model. Variant effects were computed by comparing predicted regulatory signals between the reference and alternate alleles using the variant-scoring framework implemented in AlphaGenome. The AlphaGenome generates predictions across 11 raw genomic output types based on experimental techniques such as RNA sequencing (RNA-seq), Precision Run-On coupled with cap analysis (PRO-cap), Cap Analysis of Gene Expression (CAGE), Assay for Transposase-Accessible Chromatin sequencing (ATAC-seq), DNase sequencing (DNase-seq), Transcription Factor Chromatin Immunoprecipitation Sequencing (TF ChIP-seq), Histone Chromatin Immunoprecipitation sequencing (Histone ChIP-seq), splice sites, splice junctions, splice-site usage, and contact maps. For each variant-track combination, AlphaGenome produced annotations describing the regulatory context and predicted effect size. These included variant-level information, such as variant_id and the scored genomic interval; gene-level annotations including gene_id, gene_name, gene_type, and gene_strand; assay-specific attributes including output type, assay title, track name, and track strand; and biosample metadata including biosample name, biosample type, life stage, and ontology terms. Additional regulatory descriptors, such as TF identity, histone mark type, and tissue annotations, including GTEx tissue labels, were retained where available. Quantitative outputs included raw prediction scores and normalized quantile scores, representing the magnitude and direction of variant-associated regulatory effects. All predictions were retained for downstream filtering and analysis.

### Biosample-level exploratory analysis

To characterize the distribution of AlphaGenome predictions across biological contexts, prediction outputs were summarized at the biosample level using metadata from each genomic output types (Additional file 2: Table S3). For each biosample, aggregate metrics were calculated to describe prediction abundance, strength, and regulatory breadth, including the total number of variant-track signals, the number of high-confidence signals defined by absolute quantile score ≥ 0.90, positive and negative high-confidence signal counts, the number of available genomic output types, distinct output types, represented regulatory modalities, and quantile-score summary statistics such as maximum and mean absolute quantile score (Additional file 2: Table S4). Because biosamples differed in the number of available AlphaGenome tracks, signal counts were also normalized by track availability. Normalized total and high-confidence signal burdens were calculated by dividing the corresponding signal counts by the number of available genomic output types for each biosample, providing track-adjusted estimates of prediction burden. Biosamples were first summarized across all biosample types and then filtered to retain ‘tissue biosamples’ for the main tissue-level exploratory analysis, reducing potential over-representation of cell lines or isolated cell types (Additional file 2: Table S4). Tissues were ranked by total and normalized signal burden, while regulatory breadth was assessed using the number of distinct output types and regulatory modalities. These summaries guided tissue-context selection for downstream uterus-specific analyses.

### Uterus-specific filtering and high-confidence prediction selection

Given the uterine origin of endometriosis, AlphaGenome predictions were filtered using biosample annotations. Predictions corresponding to uterus-related biosamples were retained based on metadata fields such as biosample_name, tissue annotation, and associated ontology information. AlphaGenome quantile scores were used to quantify the relative magnitude of predicted variant effects across different assays and biosamples. The quantile score represents the relative strength of a variant-induced predicted signal change compared with a background distribution of model predictions. Positive quantile scores indicate an increase in predicted signal associated with the alternate allele, whereas negative quantile scores indicate a decrease relative to the reference allele. Given the large scale and heterogeneity of the generated prediction output, a stringent high-confidence threshold was applied. Only predictions with an absolute quantile score ≥ 0.90 were retained for the main uterus-specific regulatory analyses. This threshold was used to prioritize strong predicted regulatory effects while reducing noise across the multi-modal AlphaGenome output (Fig. 1, Additional file 1 and Additional file 2: Table S5).

### Annotation of uterus-specific AlphaGenome predictions

The uterus-specific prediction table was annotated by mapping each AlphaGenome variant_id to its corresponding rsID using the hg38 SNP reference file generated during variant preprocessing (Additional file 2: Table S2). Locus information was incorporated by matching annotated rsIDs to the curated lead-SNP locus table derived from the 42 genome-wide significant endometriosis loci reported in the published GWAS study [3] (Additional file 2: Tables S6 and S7). To characterize the spatial relationship between predicted regulatory variants and their associated genes, the distance between each SNP and the corresponding gene body was calculated in base pairs using hg38 gene annotation coordinates. For each variant-gene pair, the SNP position was compared with the annotated start and end coordinates of the matched gene. Variants located within the gene body were assigned a distance of 0 bp, whereas intergenic variants were assigned the shortest linear distance to the nearest gene boundary. Additional gene-level features, including gene start, gene end, and gene strand, were incorporated into the final annotated uterus-specific prediction table (Additional file 2: Table S8).

### Regulatory category grouping and tier classification

For SNP-level downstream interpretation, uterus-specific high-confidence AlphaGenome predictions were collapsed into integrated regulatory modalities rather than analyzed as individual raw genomic output types. Although AlphaGenome generated predictions across 11 raw genomic output types, only the genomic tracks represented after uterus-specific biosample filtering and high-confidence filtering were used for tier-based classification. Accordingly, six uterus-specific regulatory modalities were defined: gene expression, promoter activity/transcription initiation, chromatin accessibility, TF binding, histone modification, and RNA splicing. RNA-seq predictions were assigned to the gene expression category, reflecting predicted transcript abundance. CAGE predictions were assigned to promoter activity, reflecting transcription start site-associated regulatory activity. ATAC-seq and DNase-seq predictions were grouped under chromatin accessibility, representing predicted open chromatin regions. TF ChIP-seq predictions were assigned to TF binding, whereas histone ChIP-seq predictions were assigned to histone modification, representing chromatin-state-associated regulatory signals. Splicing-related predictions, such as splice junctions and splice-site usage, were grouped under RNA splicing, capturing predicted effects on splice junctions and splice-site usage (Fig. 1).

Other AlphaGenome predicted genomic output types, including PRO-cap, splice sites, and contact maps, were retained in the global AlphaGenome output summary but were not present among the uterus-specific high-confidence tracks used for downstream SNP interpretation. Therefore, these outputs were not included in the uterus-specific tiering framework (Fig. 2). Tier classification was based only on the six collapsed regulatory modalities described above. Each SNP was summarized according to the number of distinct uterus-specific high-confidence regulatory modalities in which it showed predicted regulatory effects. The SNPs were then classified into five tiers according to regulatory-modality breadth: Tier 1, support across all six modalities; Tier 2, support across five modalities; Tier 3, support across four modalities; Tier 4, support across three modalities; and Tier 5, support across two or fewer modalities. This six-regulatory modality tiering framework was applied consistently to the published GWAS lead SNPs, AlphaGenome-prioritized SNPs, and outside-GWAS-lead-SNP loci (Fig. 1).

**Figure 2.**
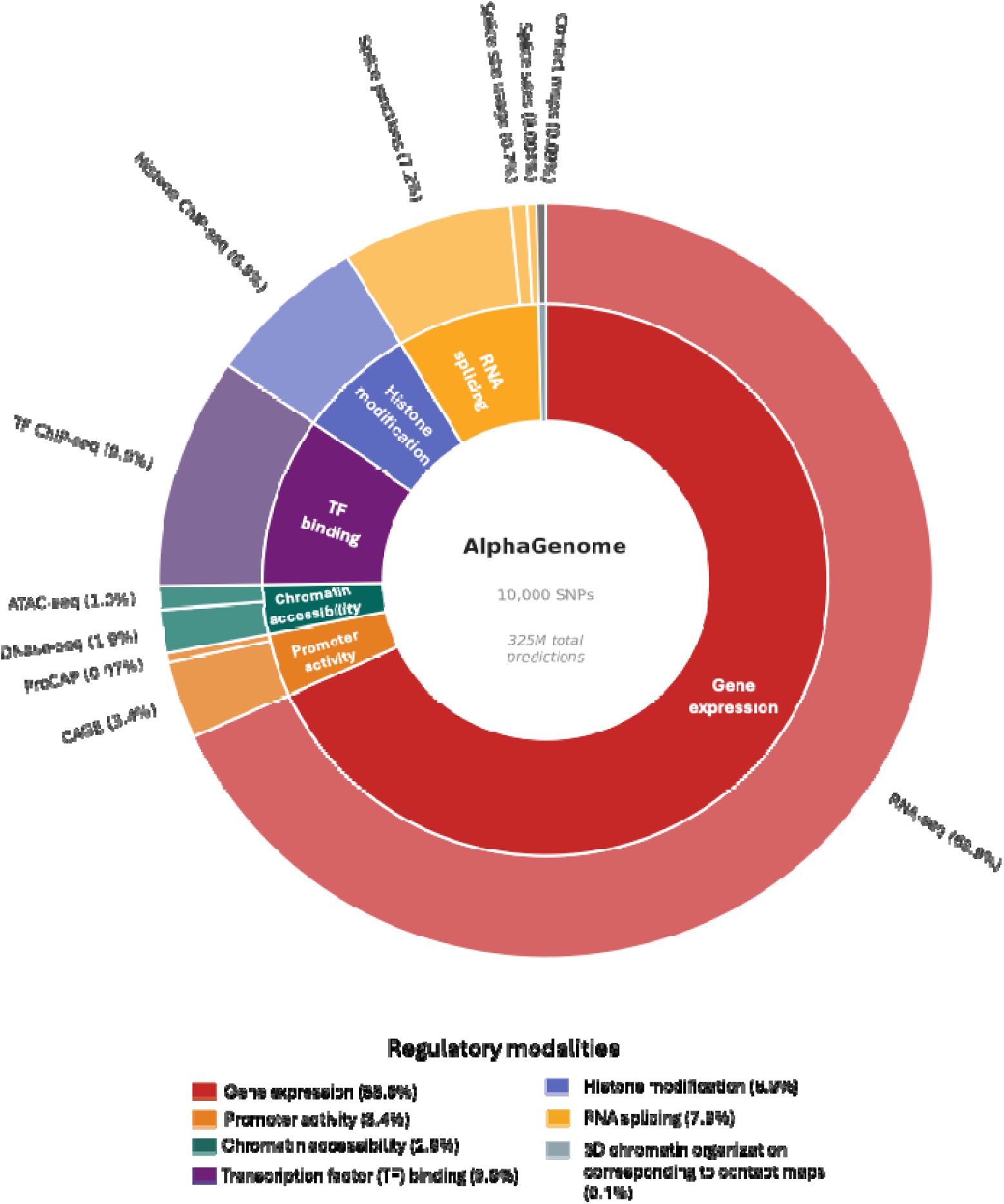
Global AlphaGenome prediction landscape across the 10,000-SNP dataset. Nested donut chart showing the total number of AlphaGenome variant-track prediction signals across 11 raw genomic output types in outer circle collapsed into seven regulatory modalities in the inner circle.

### Locus-based grouping of 42 lead GWAS SNPs and selection of AlphaGenome-prioritized representative SNPs

To compare uterus-specific AlphaGenome predictions with the published endometriosis GWAS architecture, the 42 lead SNPs were assigned to the 42 genome-wide significant loci identified in the GWAS meta-analysis [3] (Additional file 2: Table S7). In this GWAS framework, loci were defined around lead SNPs representing the most significantly associated variant within a regional association signal, using a ±500 kb window (*p* < 5 × 10^-8^). Following the same locus-based approach, the 42 published GWAS lead SNPs were used to construct ±500 kb locus windows in hg38 coordinates. All uterus-filtered SNPs with high-confidence AlphaGenome predictions were assigned to these loci based on genomic position. Where a SNP fell into overlapping locus windows, it was assigned to the nearest locus based on distance to the center of the lead-SNP region. For each locus, retained SNPs were summarized according to their AlphaGenome-predicted regulatory modalities. This included the number of unique genomic track-level predictions, the number of supported regulatory modalities, and the maximum absolute quantile score. SNP-level GWAS association *p* values were mapped back to retained SNPs using chromosome and hg19 genomic position from the original GWAS summary dataset (Additional file 2: Table S1). Within each locus, SNPs were ranked according to GWAS significance and AlphaGenome regulatory support (Additional file 2: Table S9).

To identify AlphaGenome-prioritized representative SNPs within the same GWAS loci, one alternative SNP was selected per locus using the following criteria: the SNP had to be genome-wide significant at *p* < 5 × 10 □ □, distinct from the SNP with the smallest *p* value in that locus, and supported by high-confidence uterus-specific AlphaGenome predictions. Prioritization was based first on the number of genomic track-level predictions, followed by the number of supported regulatory modalities, maximum absolute quantile score, and GWAS *p* value (Additional file 2: Table S10). These SNPs are further referred as “AlphaGenome-prioritized SNPs” (Fig. 1 and Additional file 1). The final locus-level summary table included, for each locus, the published GWAS lead SNP and the AlphaGenome-prioritized representative SNP, together with genomic position, effect and non-effect alleles, effect allele frequency, and GWAS association *p* value (Additional file 2: Table S11). Separate prediction summary tables were also generated containing the full uterus-specific AlphaGenome regulatory profiles for the published GWAS lead SNPs and AlphaGenome-prioritized SNPs representing the 42 loci (Additional file 2: Tables S12 and S13).

### Identification and characterization of outside-locus SNPs from the 10,000 endometriosis-associated GWAS SNPs

SNPs from the 10,000 GWAS-ranked variant set that did not fall within any of the 42 ±500 kb published GWAS lead SNP locus windows (n = 3,454) were classified as outside-locus or unassigned SNPs (Fig. 1, Additional file 1 and Additional file 2: Table S14). These SNPs were retained for a separate exploratory regulatory-track analysis. Genome-wide significant outside-locus SNPs were identified using the same threshold applied in the locus-based analysis, *p* < 5 × 10□□. Outside-locus SNPs were then filtered for the presence of at least one high-confidence uterus-specific AlphaGenome prediction (absolute quantile score ≥ 0.90). Each retained outside-locus SNP was summarized according to the number of supported regulatory modalities, number of unique genomic track-level predictions, maximum absolute quantile score, and GWAS *p* value. The same six-category framework and tier-classification system described above were applied to the outside-locus SNPs. SNP-level information, including rsID, genomic position, GWAS *p* value, regulatory categories, maximum absolute quantile score, and tier assignment, was recorded in the outside-locus summary (Additional file 2: Table S15).

### Linkage disequilibrium analysis between published endometriosis-associated GWAS lead SNPs and AlphaGenome-prioritized SNPs

To evaluate whether AlphaGenome-prioritized SNPs represented the same association signals as the corresponding published GWAS lead SNPs, pairwise linkage disequilibrium (LD) was calculated for each published GWAS lead SNP-AlphaGenome-prioritized SNP pair (Fig. 1 and Additional file 1). LD was estimated using the 1000 Genomes Phase 3 European reference panel to remain consistent with the ancestry-specific LD framework used in the original GWAS study. For each of the 42 loci, the corresponding published GWAS lead SNP and AlphaGenome-prioritized SNP were extracted from the European reference panel and analyzed as an SNP pair. Both r^2^ and D′ were calculated for each pair. The r^2^ value was used as the primary classification metric because it reflects how well one SNP predicts another, whereas D′ was retained as an additional LD descriptor but was not used for category assignment. SNP pairs were classified into four LD categories based on r²: strong LD, defined as r² ≥ 0.8; moderate LD, defined as 0.5 ≤ r^2^ < 0.8; weak LD, defined as 0.2 ≤ r^2^ < 0.5; and low LD, defined as r^2^ < 0.2. For summary-level interpretation, strong and moderate LD pairs were considered genetically linked to the published GWAS association signal, whereas weak and low LD pairs were considered potentially distinct or partially independent regulatory candidates within the same GWAS-defined locus. The resulting LD table included the published GWAS lead SNP, the AlphaGenome-prioritized SNP, genomic positions, r^2^, D′, and LD category for each locus (Additional file 2: Table S16).

### Cross-reference with multi-ancestry endometriosis-associated GWAS SNPs

To determine whether the outside-locus SNP set contained variants subsequently implicated in endometriosis, the outside-locus/unassigned SNP list was compared with novel lead SNPs reported in a more recent multi-ancestry GWAS study [14] (Fig. 1 and Additional file 1). Reported novel SNPs were matched against the original 10,000-SNP AlphaGenome input dataset using rsID and, where needed, chromosome-position information. For each matched SNP, locus assignment status was checked to determine whether it fell within one of the original 42±500 kb published GWAS lead SNP windows or belonged to the outside-locus/unassigned SNP group. Matched SNPs were then annotated with their AlphaGenome regulatory-category support, maximum absolute quantile score, number of unique track-level predictions, GWAS *p* value from the 10,000-SNP dataset, nearest gene reported in the new GWAS, and tier classification. The final comparison table included overlap status with the original 10,000-SNP dataset, outside-locus status, regulatory categories, and predicted uterus-specific AlphaGenome effects (Additional file 2: Table S17).

## Results

### Global AlphaGenome prediction landscape across 10,000 endometriosis-associated GWAS SNPs

AlphaGenome analysis of the 10,000 endometriosis-associated GWAS SNPs generated a total of 325,451,290 variant-track prediction signals across 11 raw regulatory output types, in all available biosamples, without restricting the initial analysis to uterus, which were further grouped into seven broader functional regulatory modalities for interpretation (Fig. 2 and Additional file 2: Table S18), including gene expression, promoter activity/transcription initiation, chromatin accessibility, TF binding, histone modification, RNA splicing, and three-dimensional chromatin organization. The landscape of global predicted genomic outputs was strongly dominated by RNA-seq, which accounted for 224,292,024 signals, corresponding to 68.9% of the total AlphaGenome output (Fig. 2). The next most represented output types were TF ChIP-seq (9.9%), splice junctions (7.2%), and histone ChIP-seq (6.9%). In contrast, CAGE (3.4%), DNase-seq (1.9%), ATAC-seq (1.0%), splice-site usage (0.7%), contact maps (0.1%), PRO-cap (0.1%), and splice sites contributed smaller proportions of the total prediction space (<0.1%) (Fig. 2 and Additional file 2: Table S18). When raw output types were collapsed into broader regulatory modalities, gene expression represented the largest component of the prediction landscape (68.9%), followed by TF binding (9.9%), RNA splicing (7.9%), histone modification (6.9%), promoter activity/transcription initiation (3.4%), chromatin accessibility (2.9%), and three-dimensional chromatin organization (0.1%) (Fig. 2).

### Uterus-specific AlphaGenome predictions and prioritization reveals regulatory heterogeneity across endometriosis GWAS loci

Biosample-level summarization of 10,000 endometriosis-associated GWAS SNPs revealed substantial variation in AlphaGenome prediction signal representation across several tissues (Additional file 2: Tables S4). The highest total number of variant-track prediction signals were observed in stomach, spleen, adrenal gland, ovary, testis, liver, sigmoid colon, kidney, spinal cord, and urinary bladder, indicating that raw signal abundance differed markedly across tissues. This variation likely reflected differences in the number and diversity of available AlphaGenome output types for each tissue. After normalization by the number of available output types, differences in signal burden across tissue remained evident, showing

Among reproductive tissues, ovary and uterus were both strongly represented (Fig. 3). Ovary contained 2,675,024 total prediction signals and 259,856 high-confidence signals across seven genomic output types, whereas uterus contained 1,432,412 total prediction signals and 147,033 high-confidence signals across eight genomic output types spanning six regulatory modalities (Fig. 4) such as gene expression (RNA-seq), promoter activity/transcription initiation associated activity (CAGE), chromatin accessibility (combined ATAC-seq and DNase-seq), histone modification (histone ChIP-seq), TF binding (TF ChIP-seq), and RNA splicing (combined splice junctions and splice-site usage) (Additional file 2: Table S19). RNA-seq was the dominant uterus-specific output type, contributing 1,132,788 predictions, followed by splice junctions, CAGE, TF ChIP-seq, histone ChIP-seq, ATAC-seq, DNase-seq, and splice-site usage.

**Figure 3.**
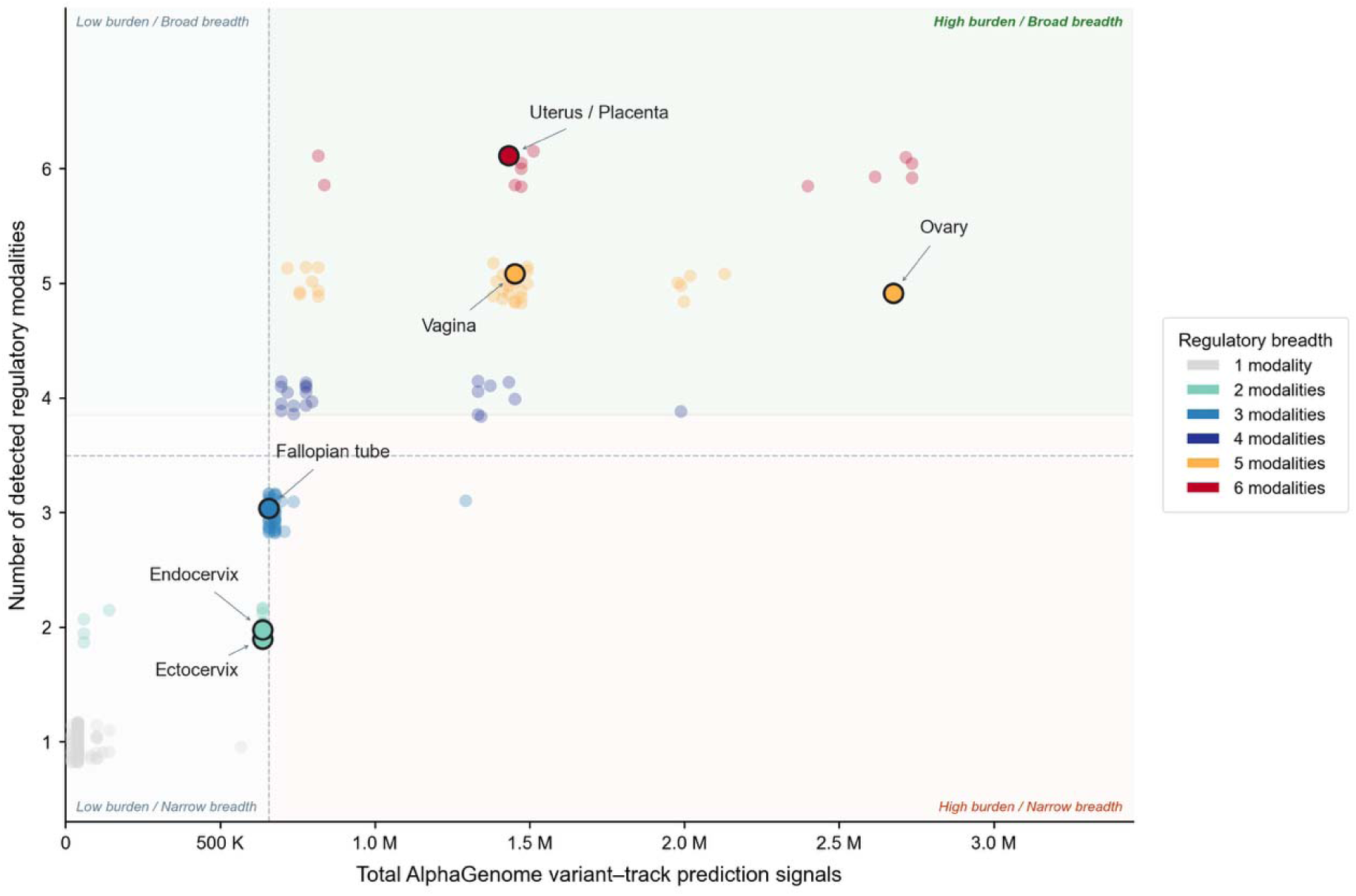
Biosample-level AlphaGenome signal burden and regulatory breadth. Each point represents a biosample plotted by the total number of AlphaGenome variant-track prediction signals for 10,000 endometriosis-associated GWAS SNPs and the number of detected regulatory modalities. Reproductive tissues are highlighted and annotated. Point color indicates regulatory breadth among the highlighted tissues, ranging from one to six detected modalities. Dashed vertical and horizontal lines define the burden and breadth thresholds used to separate biosamples into low burden/narrow breadth, low burden/broad breadth, high burden/narrow breadth, and high burden/broad breadth quadrants.

**Figure 4.**
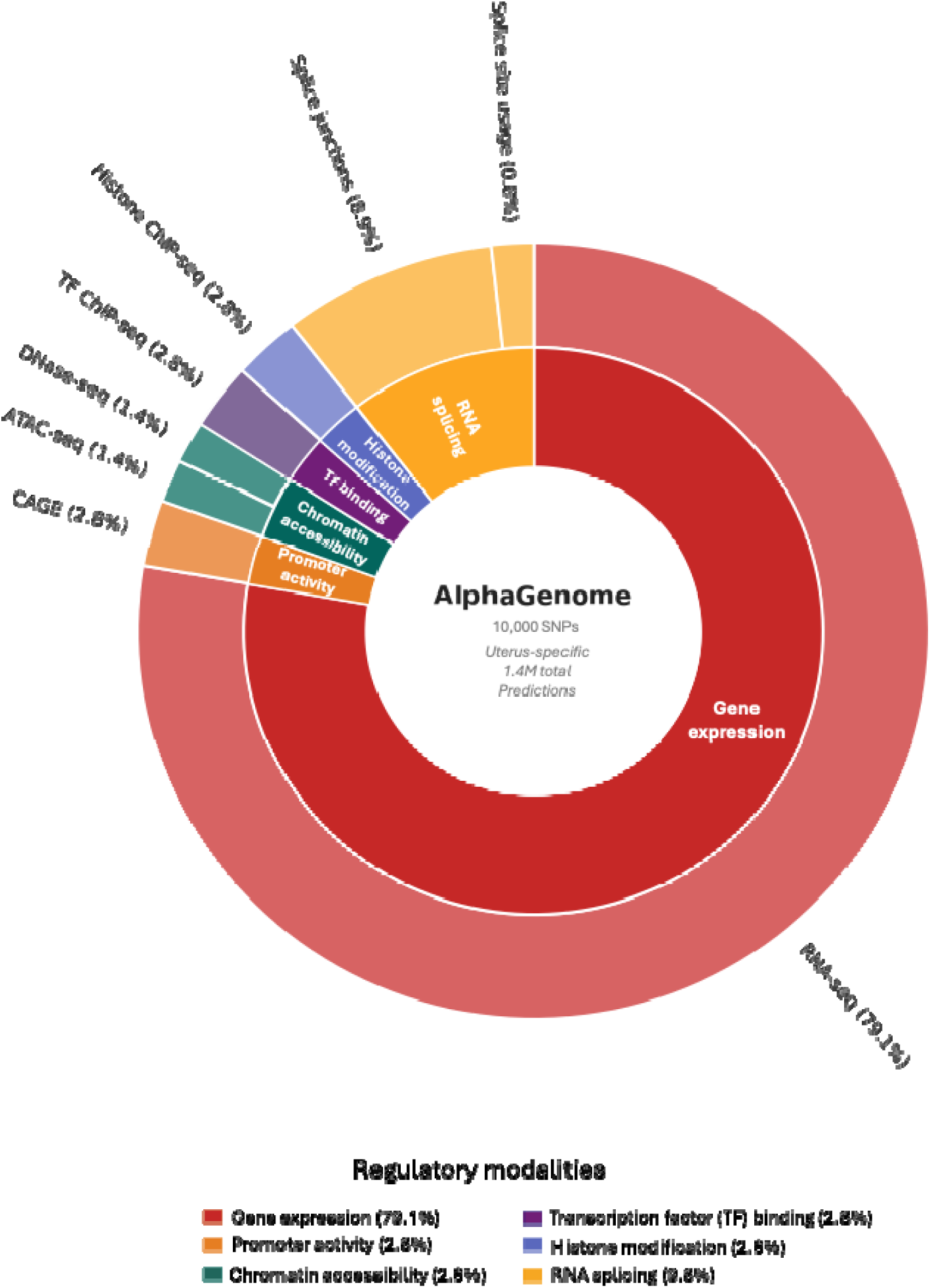
Uterus Specific AlphaGenome prediction landscape across the 10,000-SNP dataset. Nested donut chart showing the total number of AlphaGenome variant-track prediction signals across 8 raw genomic output types represented in outer circle collapsed into six regulatory modalities in inner circle.

Although ovary showed higher total and high-confidence signal counts, the uterus provided the most disease-relevant tissue context for endometriosis and captured a broader regulatory prediction landscape, with eight genomic output types compared with seven in the ovary. Therefore, the uterine context was selected for downstream locus-level analyses despite the overall signal distribution remaining largely dominated by transcript abundance–related predictions.

Moreover, following tissue-specific prioritization, uterus-filtered high-confidence SNPs were mapped to the 42 genome-wide significant endometriosis loci previously identified by GWAS meta-analysis [3]. Using regulatory prioritization strategy, AlphaGenome identified 42 alternative “AlphaGenome-prioritized SNPs” that differed from the published GWAS lead variants and were used for below comparative locus-level analysis (Fig. 5a).

**Figure 5.**
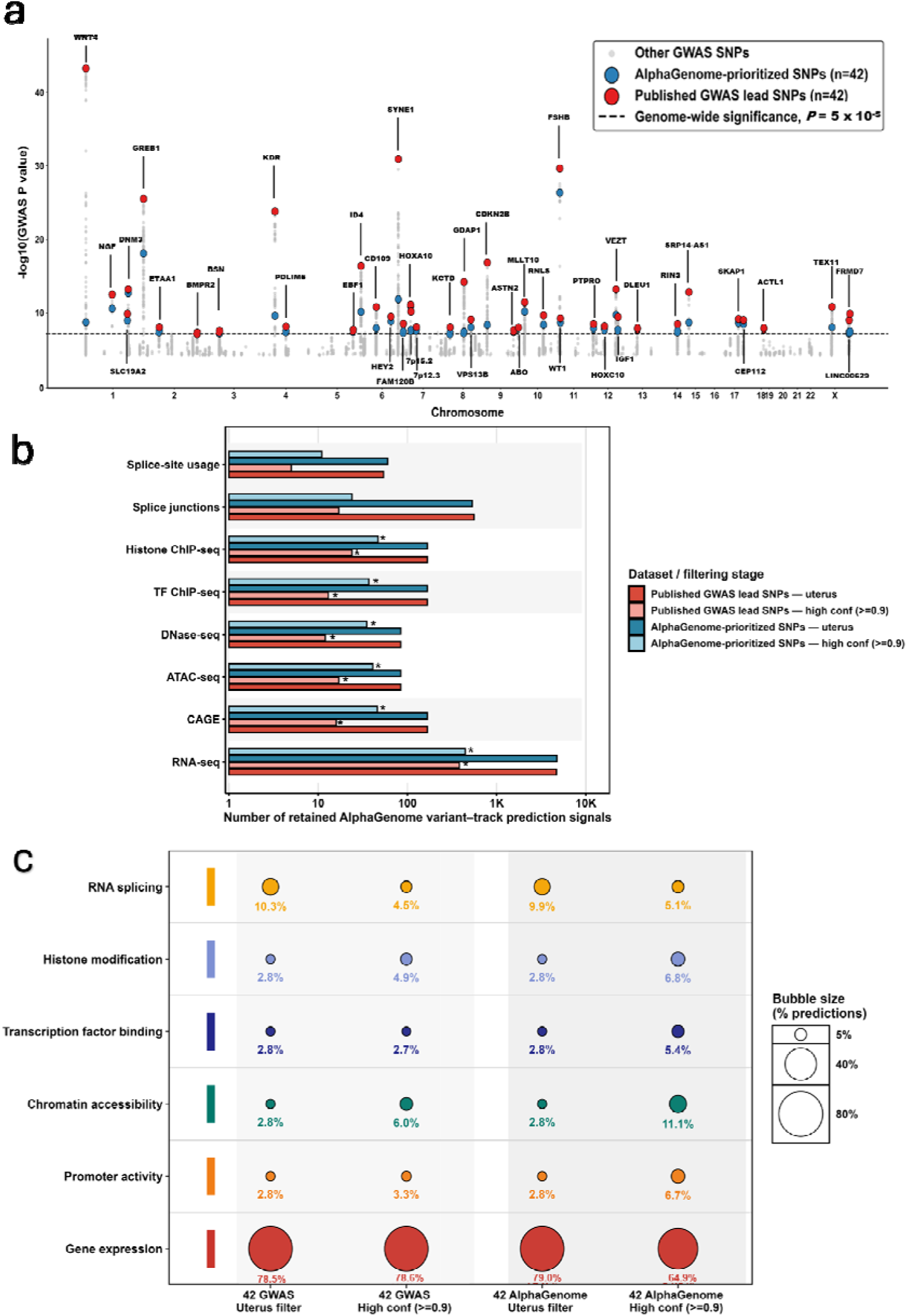
Uterus-specific AlphaGenome prioritization of endometriosis-associated SNPs. **(a)** Manhattan plot showing GWAS association strength for SNPs across chromosomes. Grey points represent other SNPs in the 10,000-SNP dataset, red points indicate the 42 published GWAS lead SNPs, and blue points indicate the 42 AlphaGenome-prioritized subthreshold SNPs selected within corresponding GWAS loci. Selected loci are labelled, and the dashed horizontal line indicates the genome-wide significance threshold of *p* = 5 × 10[[. **(b)** Absolute number of retained uterus-specific AlphaGenome variant–track prediction signals across major genomic output types for the two matched 42-SNP sets. Bars show counts for published GWAS lead SNPs after uterus tissue filtering, published GWAS lead SNPs after high-confidence filtering, AlphaGenome-prioritized SNPs after uterus tissue filtering, and AlphaGenome-prioritized SNPs after high-confidence filtering. High-confidence filtering was defined as an absolute quantile score ≥ 0.90. Asterisks indicate output types in which AlphaGenome-prioritized SNPs showed significantly higher high-confidence signal burden than published GWAS lead SNPs after FDR correction (*p* < 0.05; paired Wilcoxon signed-rank test). **(c)** Bubble plot showing the proportional distribution of retained uterus-specific prediction signals across regulatory modalities for the same two SNP sets and filtering stages. Genomic output types were collapsed into six regulatory modalities. Bubble size and percentage labels indicate the contribution of each modality to the total retained prediction signals within each dataset/filtering stage.

### Comparison of uterus-specific regulatory signal retention between GWAS lead and AlphaGenome-prioritized SNPs

Before high-confidence filtering (absolute quantile score ≥ 0.90), both the published GWAS lead SNPs and the AlphaGenome-prioritized SNPs retained predictions across the major uterus-relevant regulatory modalities with broadly comparable total signal distributions (Fig. 5b and Additional file 2: Table S19). Consistent with this pattern, paired locus-level statistical testing showed no significant differences (*p* > 0.05) between the two SNP sets across the tested modalities after false-discovery-rate (FDR) correction (Additional file 2: Table S20). This indicates that, at the level of raw uterus-filtered prediction coverage, the two matched 42-SNP sets showed similar regulatory representation.

After applying the high-confidence threshold (absolute quantile score ≥ 0.90), a clearer difference emerged between the two SNP sets. The AlphaGenome-prioritized SNPs retained more uterus-specific high-confidence regulatory signals than the published GWAS lead SNPs across most major regulatory modalities (Fig. 5b and Additional file 2: Table S19). Paired Wilcoxon signed-rank tests across the 42 matched loci showed significantly higher high-confidence signal burdens for AlphaGenome-prioritized SNPs in RNA-seq, CAGE, ATAC-seq, DNase-seq, TF ChIP-seq, and histone ChIP-seq tracks after FDR correction (*p* < 0.05) (Additional file 2: Table S20). In contrast, splice junctions and splice-site usage showed nominal differences but did not differ significantly between the two SNP sets. Although RNA-seq remained the dominant output type in both SNP sets, AlphaGenome-prioritized SNPs retained more high-confidence non-expression regulatory signals than GWAS lead SNPs, particularly for CAGE, ATAC-seq, DNase-seq, TF ChIP-seq, and histone ChIP-seq (Fig. 5b and Additional file 2: Table S19). This shift was also visible after collapsing tracks into regulatory modalities: compared with GWAS lead SNPs, AlphaGenome-prioritized SNPs showed lower relative gene expression contribution (78.6% to 64.9%) and higher contributions from promoter activity (3.3% to 6.7%), chromatin accessibility (6.0% to 11.1%), TF binding (2.7% to 5.4%), and histone modification (4.9% to 6.8%) after high-confidence filtering (Fig. 5c). These findings suggest that AlphaGenome prioritization enriched variants with stronger uterus-relevant high-confidence regulatory effects beyond gene expression alone.

### AlphaGenome-prioritized SNPs show broader uterus-specific multi-modal regulatory support than published GWAS lead SNPs

Uterus-specific AlphaGenome predictions showed that all 42 published GWAS lead SNPs and alterative 42 AlphaGenome-prioritized SNPs retained at least one high-confidence regulatory signal in the uterine context (Fig. 6a-d and Additional file 2: Table S12, S13). Tier-based stratification demonstrated substantial heterogeneity among the 42 GWAS lead SNPs, with RNA-seq representing the most consistently supported regulatory category in the uterine context (Fig. 6a). Overall, only 5 of the 42 published GWAS lead SNPs belongs to tier 1 category, while 4 additional lead SNPs were represented in tier 2 category. Most published lead SNPs were assigned to tier 4 or tier 5, indicating that many statistically defined GWAS lead SNPs showed limited predicted regulatory breadth in the uterine context (Fig. 6b). On contrary, alternative AlphaGenome-prioritized lead SNPs demonstrated substantially stronger uterus-specific regulatory support (Fig. 6c). Notably, 35 out of 42 AlphaGenome-prioritized lead SNPs were classified within tier 1-3 categories, reflecting strong regulatory support across transcriptomic, epigenomic, and splicing-related datasets in the uterus (Fig. 6d and Table 1). This indicates a broader and more integrated regulatory landscape for the AlphaGenome-prioritized SNPs compared with the GWAS lead SNPs (Additional file 2: Table S13). On the other hand, the GWAS lead SNPs exhibited limited regulatory overlap with fewer regulatory modalities predictions. Moreover, allele frequency comparisons showed no systematic difference between AlphaGenome-prioritized and published GWAS lead SNPs: prioritized SNPs had higher effect allele frequencies at 22 loci (0.426 ± 0.253) and lower at 20 loci (0.385 ± 0.223; Additional file 2: Table S11). This balance suggests that the richer regulatory signal observed in AlphaGenome-prioritized SNPs was not simply an artifact of allele frequency. Collectively, these findings suggest that AlphaGenome-based prioritization can refine GWAS loci by identifying variants with stronger predicted functional relevance in the uterine regulatory context.

**Figure 6.**
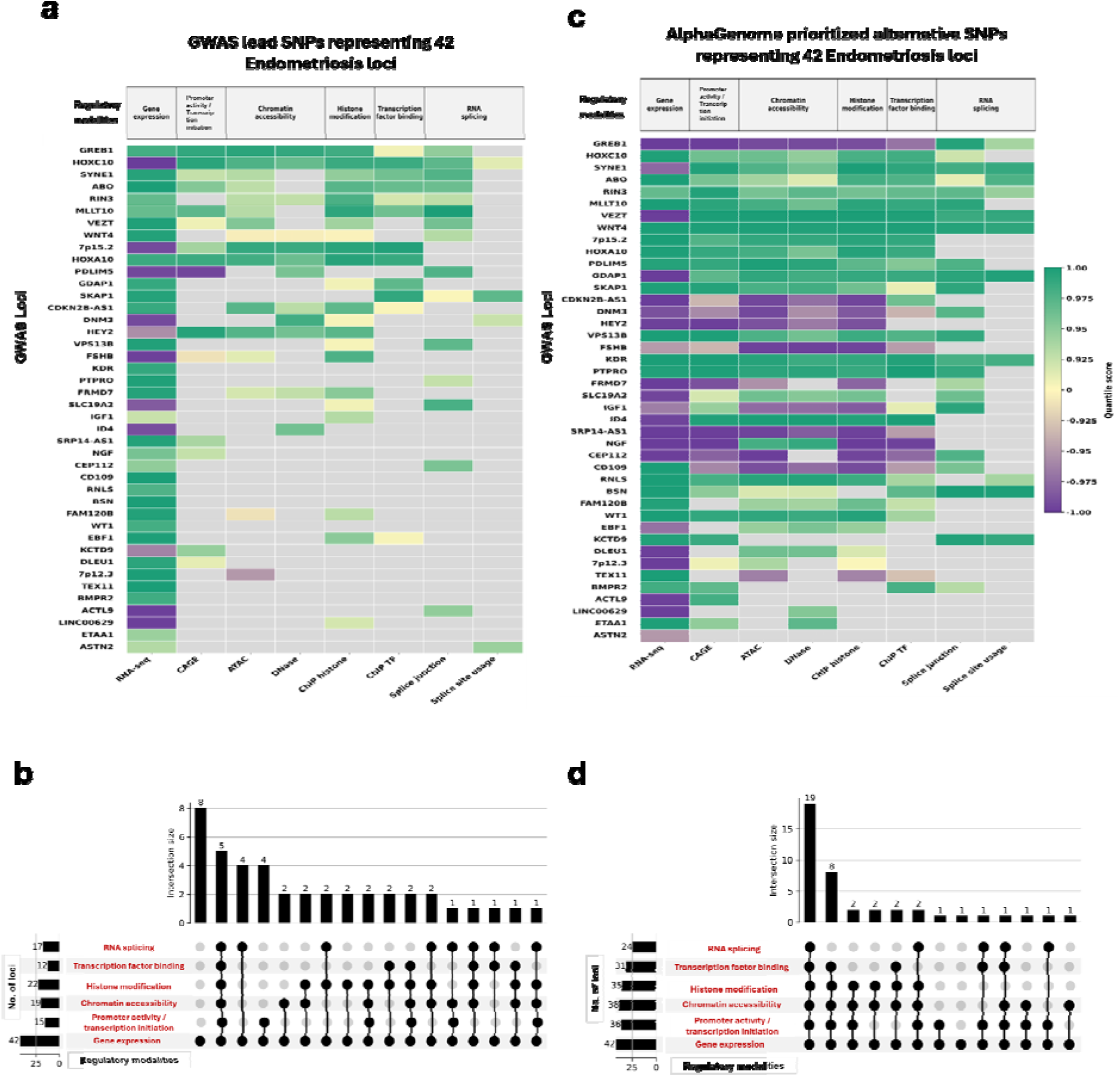
Uterus-specific regulatory profiles of GWAS lead and prioritized SNPs. (**a)** Heatmap showing high-confidence AlphaGenome predictions, defined as absolute quantile score ≥ 0.90, across six integrated regulatory modalities for the 42 published GWAS lead SNPs and the **(b)** UpSet plot showing the overlap of six integrated uterus-specific AlphaGenome regulatory modalities among the 42 published GWAS lead SNPs. **(c)** Heatmap showing high-confidence AlphaGenome predictions, defined as absolute quantile score ≥ 0.90, across six integrated regulatory modalities for the 42 AlphaGenome-prioritized representative SNPs. **(d)** UpSet plot showing the combinatorial distribution of regulatory support among the 42 AlphaGenome-prioritized SNPs selected within ±500 kb of the original GWAS lead loci. In the heatmaps **(a, c),** positive predicted effects of the alternative allele are shown in green, negative predicted effects in purple, and the absence of retained high-confidence predictions in grey. Colour intensity corresponds to the magnitude of the quantile score. In the UpSet plots **(b, d),** the six integrated regulatory categories are highlighted in red text, and bar heights represent the number of SNPs supported by each individual or combined regulatory modality.

**Table 1.** Uterus-specific regulatory tier classification of endometriosis-associated loci. Legends: Loci were stratified according to the number of independent regulatory modalities supported by uterus-specific AlphaGenome predictions. Six regulatory modalities were considered: gene expression (RNA-seq), promoter activity (CAGE), chromatin accessibility (ATAC-seq and/or DNase-seq), histone modification (histone ChIP-seq), transcription factor binding (TF ChIP-seq), and RNA splicing (splice junction and/or splice-site usage). For each locus, the presence of at least one high-confidence signal, defined as absolute quantile score ≥ 0.90, within a regulatory modality was counted as one unit of evidence. Each regulatory modality was counted once per locus, regardless of the number of individual tracks, genes, or transcripts detected within that category. Loci were classified into five tiers based on total regulatory support: Tier 1, six categories; Tier 2, five categories; Tier 3, four categories; Tier 4, three categories; and Tier 5, two or fewer categories. Higher-tier loci represent variants with broader multi-layer regulatory support across transcriptional, chromatin-associated, and post-transcriptional processes. Asterisks indicate loci that were previously resolved by GWAS fine-mapping as high-confidence candidate causal loci and were also identified in the present AlphaGenome analysis.

| Tier | Number of regulatory modalities | AlphaGenome predictions for the 42 lead SNPs representing the GWAS loci |  | Representative loci from AlphaGenome-selected SNPs |  |
| --- | --- | --- | --- | --- | --- |
|  |  | Number of loci | Loci | Number of loci | Loci |
| Tier 1 | 6 | 5 | *GREB1*/2p25.1, *SYNE1*/6q25.1*, *ABO*/9q34.2, *MLLT10*/10p12.31, *HOXC10*/12p13.13* | 19 | *ABO*/9q34.2, *CD109*/6q13, *CEP112*/17q24.1, *DNM3*/1q24.3, *GDAP1*/8q21.11, *GREB1*/2p25.1, *HOXC10*/12p13.13*, *IGF1*/12q23.2, *KDR*/4q12, *MLLT10*/10p12.31, *PDLIM5*/4q22.3, *PTPRO*/12p12.3, *RIN3*/14q32.12, *RNLS*/10q23.31, *SKAP1*/17q21.32, *SYNE1*/6q25.1*, *VEZT*/12q22, *VPS13B*/8q22.2, *WNT4*/1p36.12 |
| Tier 2 | 5 | 4 | 7p15.2/7p15.2, *HOXA10*/7p15.2*, *RIN3*/14q32.12, *VEZT*/12q22 | 11 | 7p15.2/7p15.2, *BSN*/3p21.31, *CDKN2B-AS1*/9p21.3, *FRMD7*/Xq26.2, *FSHB*/11p14.1, *HOXA10*/7p15.2*, *ID4*/6p22.3, *NGF*/1p13.2, *SLC19A2*/1q24.2, *SRP14-AS1*/15q15.1, and *WT1*/11p14.1 |
| Tier 3 | 4 | 4 | *PDLIM5*/4q22.3, *HEY2*/6q22.31, *CDKN2B-AS1*/9p21.3, *FSHB*/11p14.1 | 5 | 7p12.3/7p12.3, *BMPR2*/2q33.1, *FAM120B*/6q27, *HEY2*/6q22.31, |
|  |  |  |  |  | *TEX11/Xq13.1* |
| Tier 4 | 3 | 9 | *EBF1/5q33.3,*<br>*FAM120B/6q27,*<br>*FRMD7/Xq26.2,*<br>*GDAP1/8q21.11,*<br>*KCTD9/8p21.2,*<br>*SKAP1/17q21.32,*<br>*SLC19A2/1q24.2,*<br>*VPS13B/8q22.2,*<br>*WNT4/1p36.12* | 4 | *DLEU1/13q14.2,*<br>*EBF1/5q33.3,*<br>*ETAA1/2p14,*<br>*KCTD9/8p21.2* |
| Tier 5 | 2 | 14 | *7p12.3/7p12.3,*<br>*ACTL9/19p13.2,*<br>*CD109/6q13,*<br>*CEP112/17q24.1,*<br>*DLEU1/13q14.2,*<br>*DNM3/1q24.3,*<br>*ETAA1/2p14,*<br>*ID4/6p22.3,*<br>*IGF1/12q23.2,*<br>*KDR/4q12,*<br>*LINC00629/Xq26.3\*,*<br>*NGF/1p13.2,*<br>*PTPRO/12p12.3,*<br>*SRP14-AS1/15q15.1* | 2 | *ACTL9/19p13.2,*<br>*LINC00629/Xq26.3\** |
|  | 1 | 6 | *ASTN2/9q33.1,*<br>*BMPR2/2q33.1,*<br>*BSN/3p21.31,*<br>*RNLS/10q23.31,*<br>*TEX11/Xq13.1,*<br>*WT1/11p14.1* | 1 | *ASTN2/9q33.1* |

### AlphaGenome-prioritized SNPs are mostly linked to GWAS lead SNPs, while 14 loci show potential independent regulatory candidates for endometriosis

LD analysis identified 14 loci in which the AlphaGenome-prioritized SNP showed weak-to-low LD with the corresponding published GWAS lead SNP, defined as r² < 0.5 (Additional file 2: Table S16). These included nine loci with moderate LD, defined as 0.2 ≤ r² < 0.5: *CD109*/6q13, *ETAA1*/2p14, *GDAP1*/8q21.11, *KDR*/4q12, *RIN3*/14q32.12, *SLC19A2*/1q24.2, *SRP14-AS1*/15q15.1, *TEX11*/Xq13.1, and *VEZT*/12q22. Five additional loci showed low LD, defined as r² < 0.2: *SYNE1*/6q25.1, *CDKN2B-AS1*/9p21.3, *LINC00629*/Xq26.3, *GREB1*/2p25.1, and *WNT4*/1p36.12. These 14 weak-to-low LD loci may represent distinct or partially independent regulatory candidates within the same GWAS-defined regions. Importantly, among these, eight AlphaGenome-prioritized SNPs were classified as tier 1, three as tier 2, and one each as tier 3, tier 4, and tier 5, showing high-confidence support across all six integrated uterus-specific regulatory modalities.

In contrast, the remaining 28 of 42 loci showed at least moderate LD, defined as r² ≥ 0.5, between the published GWAS lead SNP and the AlphaGenome-prioritized SNP. Of these, 22 loci showed strong LD, defined as r² ≥ 0.8, whereas six loci showed moderate LD, defined as 0.5 ≤ r² < 0.8. Among these 28 strong-or moderate-LD loci, 11 AlphaGenome-prioritized SNPs were classified as tier 1, eight as tier 2, four as tier 3, three as tier 4, and two as tier 5. These results indicate that, in most loci, AlphaGenome prioritization identified SNPs that remained genetically linked to the original GWAS association signal while providing stronger predicted uterus-specific regulatory support. At the same time, several low-LD prioritized SNPs also showed broad regulatory support, suggesting potential additional regulatory candidates within the same GWAS-defined regions. The complete list of published GWAS lead SNPs, AlphaGenome-prioritized SNPs, r² values, D′ values, and LD interpretation and tier-based categories is provided in Additional file 2: table S16.

### Outside-locus analysis identifies 167 additional genome-wide significant SNPs with uterus-specific regulatory support

Among SNPs located outside the 42 published GWAS lead SNP locus windows, 167 SNPs reached genome-wide significance (*p* < 5 × 10 □ □) and showed high-confidence AlphaGenome predictions in at least one uterus-specific regulatory modality (Additional file 2: Table S14-S15). Of these outside-locus SNPs, six were classified as tier 1, including rs1482061, rs7772579, rs6557140, rs2982571, rs79626929, and rs12631337. Five of these SNPs mapped to chromosome 6 within the 6.q25.1 cluster (hg38) which spans a regulatory block including genes such as *SYNE1, ESR1*, *CCDC170*, *AKAP12*, *ZBTB2*, *ARMT1*, and *RMND1* (Fig. 7a and Additional file 2: Table S21). Notably, these variants were not captured within the predefined 42 GWAS lead SNP locus windows yet exhibited tier 1 regulatory support in the uterine context, suggesting that the AlphaGenome-based prioritization identifies additional high-confidence regulatory variants within known disease-relevant genomic regions that are not recovered by conventional locus-based GWAS lead SNP definitions.

**Figure 7.**
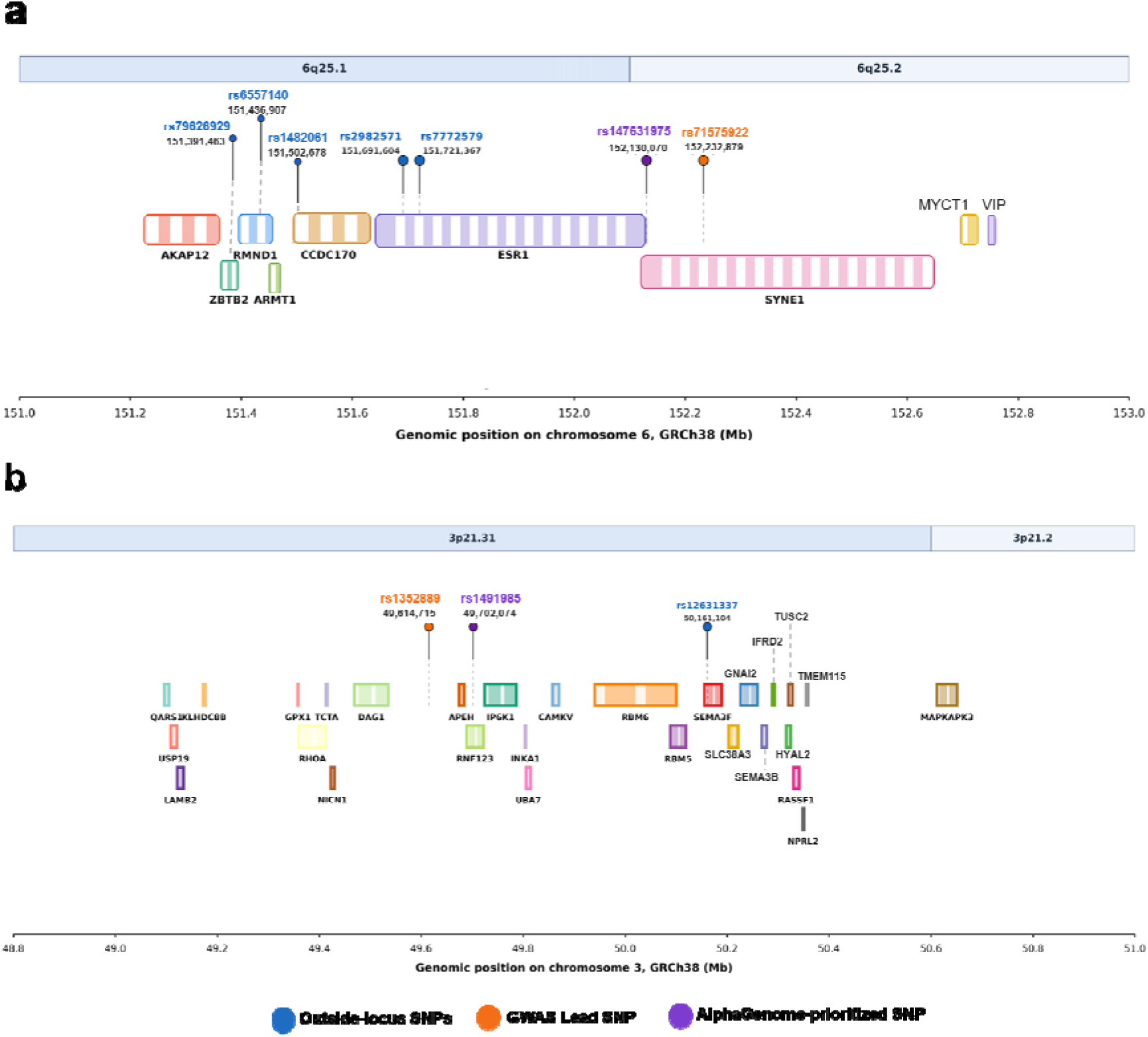
Genomic positions of endometriosis-associated SNPs on chromosomes 6 and 3. Schematic representation of selected endometriosis-associated genomic loci showing GWAS lead SNPs, AlphaGenome-prioritized SNPs, outside-locus SNPs, and nearby Ensembl-annotated AlphaGenome predicted protein coding genes using GRCh38 coordinates. (a) Chromosome 6q25.1–6q25.2 locus containing the *ESR1/SYNE1* region. (b) Chromosome 3p21.31–3p21.2 locus containing genes surrounding the selected chromosome 3 SNPs. Outside-locus SNPs are shown in blue, the GWAS lead SNPs are shown in orange, and the AlphaGenome-prioritized SNPs are shown in purple. Gene boxes indicate Ensembl-annotated genes, and genomic positions are shown in megabases according to GRCh38.

The remaining tier 1 SNP out of six outside-locus SNPs (rs12631337) mapped to chromosome 3 at chr3:50,161,104 in hg38 coordinates and was located within a gene-rich region containing multiple protein-coding genes, including *SEMA3F*, *GNAI2*, *HYAL2*, *RBM5*, *RBM6*, *TUSC2*, *UBA7*, *TMEM115*, *IP6K1*, *APEH*, *SEMA3B*, *RASSF1*, *IFRD2*, *SLC38A3*, *MAPKAPK3*, and *NPRL2* (Fig. 7b and Additional file 2: Table S21). Other genome-wide significant outside-locus SNPs were distributed across lower regulatory tiers, with 9 SNPs classified as tier 2, the 14 SNPs as tier 3, the 38 SNPs as tier 4, and 100 SNPs as tier 5. Within tier 5, 51 SNPs showed support across two regulatory modalities, whereas 49 SNPs retained only one high-confidence regulatory modality. Thus, although most genome-wide significant outside-locus SNPs showed narrower regulatory support, a smaller subset displayed broad multi-layer uterus-specific regulatory evidence. These findings indicate that, beyond the predefined published GWAS loci, other genome-wide significant variants may carry uterus-specific regulatory information relevant for downstream functional prioritization (Additional file 2: Table S14-S21).

### Cross-reference with multi-ancestry GWAS SNPs reveals high-confidence uterus-specific regulatory support for newly reported endometriosis loci

To assess whether uterus-specific AlphaGenome predictions supported recently reported endometriosis-associated variants, the AlphaGenome-ranked 10,000-SNP dataset was cross-referenced with 37 novel lead SNPs reported in a recent GWAS study [4,5]. Of these, 22 variants were present in the original 10,000-SNP dataset, whereas 15 were absent (Additional file 2: Table S17). All 22 overlapping SNPs were located outside the original 42 published endometriosis GWAS loci, based on the ±500 kb lead-SNP window definition, and none corresponded to the original GWAS lead SNPs. The overlapping SNPs showed variable degrees of uterus-specific regulatory support. Based on the number of high-confidence AlphaGenome regulatory modalities detected, one SNP was classified as tier 1, three as tier 2, four as tier 3, five as tier 4, and nine as tier 5 (Additional file 2: Table S17). Notably, 13 of the 22 overlapping SNPs belonged to tiers 1-4, indicating support across three to six regulatory modalities. The strongest signal was observed for rs1799293, reported near *ARHGAP26*, which showed high-confidence predictions across all six integrated categories, tier 1 (Fig. 8).

**Figure 8.**
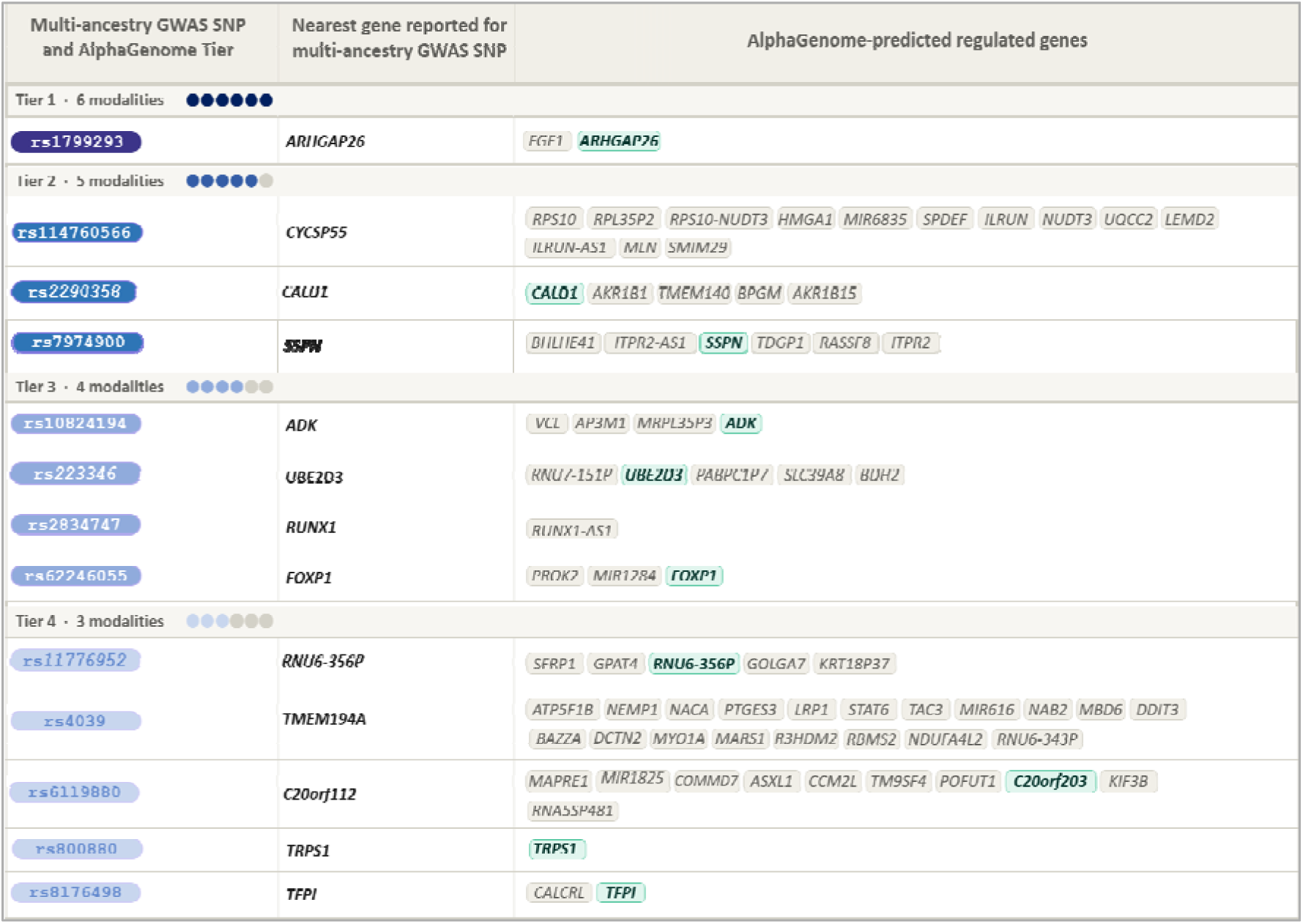
Cross-reference of multi-ancestry endometriosis GWAS SNPs with uterus-specific AlphaGenome-predicted regulated genes. Multi-ancestry endometriosis GWAS SNPs overlapping the top 10,000 AlphaGenome-ranked variants are categorized into tier 1-4 according to the number of supported high-confidence regulatory modalities. The left column shows the SNP identifier and AlphaGenome tier. The middle column shows the nearest gene for the SNPs in the multi-ancestry GWAS, and the right column shows genes or transcripts predicted by AlphaGenome to be regulated in the uterus-specific dataset. Highlighted genes indicate common genes identified both as multi-ancestry GWAS-reported genes and as AlphaGenome-predicted regulated targets.

Importantly, several genes reported as nearest genes in the recent GWAS were also detected as AlphaGenome-predicted regulated genes in the uterus-specific output, supporting concordance between the external GWAS annotation and the present regulatory predictions (Fig. 8). For example, rs2290358 was reported near *CALD1*, and AlphaGenome also predicted regulation of *CALD1*, together with additional transcripts including *AKR1B1, TMEM140, BPGM, AKR1B15*, and nearby unannotated Ensembl transcripts. Similarly, rs1799293 was reported near *ARHGAP26* and showed AlphaGenome-predicted effects involving *ARHGAP26* as well as *FGF1*. The rs7974900 variant, reported near the SSPN region, was also linked to predicted regulation of SSPN and additional transcripts including *BHLHE41, ITPR2,* and *RASSF8*. Other examples included rs223346 near *UBE2D3*, rs62246055 near *FOXP1*, rs10824194 near *ADK*, and rs2834747 near *RUNX1*, for which the GWAS-reported nearby gene was also represented among the AlphaGenome-predicted regulated genes or transcripts (Fig. 8 and Additional file 2: Table S22). Together, this cross-reference indicates that a subset of recently reported multi-ancestry endometriosis GWAS SNPs already present in the 10,000-SNP AlphaGenome dataset showed high-confidence uterus-specific regulatory evidence. In several cases, AlphaGenome not only recovered the GWAS-reported nearby gene but also identified additional potentially regulated transcripts and multilayer regulatory modalities, providing functional support for these newly reported endometriosis-associated loci.

## Discussion

Encyclopedia of DNA Elements (ENCODE) project data and subsequent Mendelian randomization studies have consistently shown that ∼90% of GWAS signals across complex diseases map to non-coding functional elements [31,32]. Endometriosis is not an exception. The majority of GWAS signals identified for endometriosis map to non-coding or intergenic regions of the genome [33,34] suggesting that these variants confer disease risk through regulatory rather than protein-altering mechanisms [35]. However, functional interpretation of these loci remains challenging because the lead SNP at a GWAS locus selected based on statistical association strength rather than functional evidence, may not be necessarily the causal or most functionally relevant variant. In regions of LD, the lead associated SNP may act as a proxy for nearby variants with stronger functional effects on gene regulation. Moreover, assigning GWAS loci to causal genes remains challenging because the functionally affected gene is not always the nearest gene to the associated variant. This is particularly relevant for non-coding risk variants, whose functional impact and target genes are often difficult to define [36], because such variants may influence distal regulatory elements, chromatin interactions, or non-coding RNA-mediated regulatory networks.

Endometriosis, being a heterogenous disorder involving multiple molecular mechanisms, is also characterized by dysregulated non-coding RNA networks, including miRNAs, lncRNAs, and circRNAs in endometriotic lesions, eutopic endometrium, and peripheral immune compartments [37–39]. These regulatory networks interact with epigenetic modifications, chromatin remodeling, and TF activity to coordinate tissue-specific gene expression. Importantly, because many non-coding RNA loci and regulatory elements reside within non-coding genomic regions, GWAS SNPs mapping to these regions may contribute to disease susceptibility through disruption of regulatory and post-transcriptional mechanisms that are not captured by protein-coding-centric analyses. Consequently, integrative computational frameworks capable of modeling long-range regulatory effects are increasingly required for functional interpretation of disease-associated non-coding variants.

Within this context, application of the AlphaGenome framework [26] enabled systematic functional annotation of 10,000 endometriosis-associated GWAS SNPs in a uterus-specific regulatory landscape. By leveraging up to 1 Mb of surrounding genomic sequence context for each variant, AlphaGenome facilitated evaluation of variant effects across multiple regulatory modalities, including gene expression, promoter activity, chromatin accessibility, TF binding, histone modification, and RNA splicing. We used a tier-based classification framework for integration of these predictions which further enabled refinement of uterus-specific regulatory prioritization across GWAS loci. Collectively, these findings support the utility of long-range sequence-based modeling approaches for improving identification and prioritization of potentially functional non-coding variants underlying endometriosis susceptibility.

Previous GWAS follow-up studies have largely relied on experimentally derived functional annotation resources, including expression quantitative trait locus datasets such as GTEx, regulatory annotations from ENCODE and related epigenomic consortia, fine-mapping, Mendelian randomization, and computational causal-gene prioritization methods [40]. More recent network-based approaches, such as SigNet, integrate within-locus evidence with cross-locus gene and regulatory network information to prioritize likely causal genes at GWAS loci [36]. Although these approaches are valuable for moving from association signals toward candidate genes, they remain constrained by the availability, resolution, and tissue relevance of the underlying annotation datasets, particularly for less studied diseases like endometriosis. In particular, many resources have incomplete coverage of disease-relevant uterine context and often prioritize genes rather than directly modelling variant-level regulatory effects across multiple molecular layers. In contrast, our analysis enabled integrated evaluation of endometriosis-associated variants across multiple predicted regulatory layers within a uterus-specific context. This provides a complementary framework for interpreting non-coding GWAS signals beyond statistical association alone.

The comparison between published GWAS lead SNPs [3] and AlphaGenome-prioritized SNPs showed that all published GWAS lead SNPs retained at least one predicted regulatory signal in the uterus-specific analysis, indicating that the published GWAS loci remain functionally relevant. This was consistent with partial concordance with independent fine-mapping evidence reported by Rahmioglu *et al*. [3], where four of six high-confidence non-coding variants also showed multi-layer regulatory support in the present analysis. However, AlphaGenome-based prioritization identified alternative SNPs within the same GWAS-defined regions that exhibited broader multimodal regulatory support, suggesting improved functional resolution beyond statistical association alone. Notably, only five published GWAS lead SNPs were classified as tier 1, whereas 19 AlphaGenome-prioritized SNPs were assigned to tier 1, reflecting a substantially higher proportion of variants with convergent support across all six regulatory categories following re-prioritization. These results are consistent with previous work showing that LD can make it difficult to distinguish lead associated variants from nearby causal or functionally relevant variants within the same GWAS locus [40].

A subset of loci showed low LD between published GWAS lead SNPs (42 loci) and AlphaGenome-prioritized SNPs, indicating that the most statistically significant GWAS SNPs do not always correspond to the strongest predicted regulatory effect. Notably, among 14 loci with weak-to-low LD (r² < 0.5), eight loci (*RIN3*/14q32.12, *GDAP1*/8q21.11, *KDR*/4q12, *VEZT*/12q22, *CD109*/6q13, *WNT4*/1p36.12, *GREB1*/2p25.1, *SYNE1/*6q25.1) were classified as tier 1 with six regulatory modalities. Collectively, these findings highlight the potential of regulatory annotation to refine causal inference and identify potential functional variation beyond standard LD-based interpretation of GWAS signals.

Among these prioritized tier 1 loci, the *WNT4*/1p36.12,*VEZT*/12q22 and KDR/4q12 loci are of particular relevance, as they have been consistently identified among the most significant SNPs associated with endometriosis [41–47]. These loci also showed weak-to-low LD (r² < 0.5) between the AlphaGenome-prioritized SNPs and the corresponding GWAS lead variants, suggesting that the strongest association signals do not necessarily reflect the most likely causal regulatory variants. Together, these features provide informative examples of how regulatory annotation can refine causal variant prioritization beyond published GWAS lead SNPs. The *WNT4*/1p36.12 contains a high-affinity estrogen receptor alpha-binding site at the locus, suggesting direct hormonal regulation of its expression [48]. WNT4 protein expression is reduced in both eutopic and ectopic endometrium in endometriosis compared to normal endometrium [49], it also regulates stromal cell proliferation, apoptosis, and decidualization, processes essential for endometrial tissue remodelling. These alterations may disrupt endometrial homeostasis and promote aberrant tissue remodelling, contributing to the development and persistence of endometriotic lesions. Moreover, the expression of *WNT4* in normal peritoneum suggests that endometriosis arising via metaplasia cannot be completely excluded [50]. Similarly, the *VEZT*/12q22 locus is characterised by increased *VEZT* expression in ectopic compared with eutopic endometrium in women with endometriosis [51], and its mRNA and protein expression shows a menstrual cycle stage-specific increase in endometrial glands during the secretory phase [52]. The *VEZT*/12q22 locus contains an NF-κB binding site, suggesting that risk variants may influence inflammatory signaling pathways in endometriosis through modulation of NF-κB–mediated transcriptional regulation [52]. The *KDR*/4q12 locus encodes *VEGFR2 gene*, the principal mediator of VEGF-driven angiogenesis and endothelial proliferation [47]. The risk variant (rs17773813[G]) at *KDR/*4q12 locus is associated with disease severity, with larger effect sizes in stage III/IV relative to stage I/II endometriosis [47] [3]. Collectively, these loci represent functionally coherent examples spanning hormonal regulation (*WNT4*/1p36.12), inflammatory signalling (*VEZT*/12q22), and angiogenesis (*KDR*/4q12) involved in endometriosis related pathways. Thus, at well-established endometriosis susceptibility loci, AlphaGenome consistently prioritized alternative SNPs, classifying them as tier 1 across all six regulatory modalities. Notably, these SNPs were absent from the original GWAS reports and showed weak-to-low LD (r² < 0.5) with the corresponding lead variants, underscoring the potential of regulatory annotation to refine causal variant identification beyond conventional GWAS-based approaches.

The alternate SNP was also suggested at the *GREB1/*2p25.1 locus, which was likewise among the eight tier 1 loci showing weak-to-low LD with the published GWAS lead SNP, and encodes an early estrogen-responsive gene and a key co-regulator of estrogen receptor activity in hormone-driven tissues. Increased *GREB1* expression has also been reported in peritoneal endometriotic lesions compared with eutopic endometrium, suggesting a role in the estrogen-driven proliferative environment of ectopic lesions [53]. Similarly, the *SYNE1*/6q25.2 locus, another low-LD tier 1 locus identified by AlphaGenome, encompassing *ESR1*, is one of the most architecturally complex susceptibility regions in endometriosis genetics. This locus is regulated through long-range chromatin interactions involving *ESR1, CCDC170, ARMT1*, and *SYNE1* genes, with distal regulatory elements and transcription factors coordinating *ESR1* expression [54]. This suggests that intronic variants in *SYNE1*/6q25.2 locus may function as non-coding regulatory elements rather than protein-altering variants. Consistently, the multimodal regulatory activity identified at the *SYNE1*/6q25.2 locus by AlphaGenome supports the functional relevance of this region in estrogen-dependent endometriosis pathogenesis.

Finally, the CD109/6q13 locus, also belonging to the subset of eight tier 1 loci in which the AlphaGenome-prioritized SNP exhibited weak-to-low LD with the published GWAS lead variant, encodes a GPI-anchored glycoprotein that negatively regulates TGF-β signalling [55,56]. Given the well-established involvement of TGF-β signalling in endometriosis pathogenesis including inflammation, epithelial-mesenchymal transition, angiogenesis, and fibrosis [57], CD109 represents a key modulator of these processes. Hence these AlphaGenome-prioritized tier 1 loci, particularly those showing low LD with published GWAS lead SNPs, represent examples of improved functional prioritization beyond statistical association alone and further support the contribution of regulatory mechanisms to endometriosis susceptibility.

Another important finding in our study was the outside-locus analysis, which extended the AlphaGenome interpretation beyond the published 42 GWAS lead SNP windows. Although the primary analysis was restricted to SNPs located within ±500 kb of each lead SNP, 3,454 SNPs lay outside these predefined regions, including 167 genome-wide significant SNPs with high-confidence uterus-specific regulatory predictions. While most of these variants showed limited regulatory support, six SNPs (rs1482061, rs7772579, rs6557140, rs2982571, rs79626929, and rs12631337) were classified as tier 1 variants. Five of these SNPs (rs1482061, rs7772579, rs6557140, rs2982571, and rs79626929) are located on chromosome 6 and are predicted to regulate genes within *ESR1*/6q25.1 and *SYNE1*/6q25.2 loci including *ESR1, SYNE1, AKAP12, ZBTB2, ARMT1* and *RMND1* (Fig. 7a). In our study, these variants are designated as outside-locus SNPs because they fall outside the 500 kb window used for locus definition, despite being in proximity to the published GWAS lead SNP rs71575922 and the AlphaGenome-prioritized SNP rs147631975 regulating *SYNE1* gene. Notably, these variants cluster near the outside-locus tier 1 SNPs, suggesting the presence of an extended regulatory region spanning the *ESR1*/6q25.1-*SYNE1*/6q25.2 locus.

Among the outside-locus SNPs, tier 1 variant rs7772579 is in strong LD (r² = 0.98) with the previously reported endometriosis-associated variant rs2206949 in *ESR1* gene [46]. Additionally, tier 2 SNP rs1971256, located in *CCDC170* has also been reported as an endometriosis risk variant [46]. The identification of multiple prioritized variants within *ESR1*/6q25.1-*SYNE1*/6q25.2 region, further supports that this is a robust and consistently replicated endometriosis susceptibility loci. *ESR1* gene, encoding estrogen receptor α showed robust genome-wide significant association across multiple meta-analyses, with several independent signals at this locus further substantiating its central role in estrogen-driven lesion growth and inflammation [8,46]. Nearby genes, including *SYNE1, CCDC170*, *AKAP12*, *ZBTB2*, *ARMT1*, and *RMND1* have also been implicated through GWAS and transcriptome-wide analyses, showing expression correlations with *ESR1* and cell-specific expression in endometrial epithelial cells during the secretory phase, implicated in implantation impairment, often associated with endometriosis [8,46,58]. *AKAP12*, while not mapped to published GWAS lead SNPs, showed negative correlation with *ESR1* and functions in cytoskeletal and signalling regulation, supporting its potential modulatory role within this network [59].

Similarly, tier 1 outside-locus SNP rs12631337 predicted to regulate tumor suppressor gene cluster on chromosome 3, including *SEMA3F*, *GNAI2*, *HYAL2*, *RBM5*, *RBM6*, *TUSC2*, *UBA7*, *TMEM115*, *IP6K1*, *APEH*, *SEMA3B*, *RASSF1*, *IFRD2*, *SLC38A3*, *MAPKAPK3*, and *NPRL2* (Fig. 7b). This SNP is in proximity to the *BSN*/3p21.31 locus together with the GWAS lead SNP rs1352889. However, rs12631337 falls outside the predefined ±500 kb locus boundary and was therefore classified as an outside-locus variant despite its likely regulatory connection to this region. Among the genes predicted to be regulated by this SNP, *RASSF1* is particularly notable because its tumor-suppressor isoform, *RASSF1A*, has been reported to show consistent epigenetic dysregulation in endometriosis, including promoter hypermethylation in both ectopic and eutopic endometrium, reduced expression, and association with disease severity [60].

These findings indicate that additional genome-wide significant variants outside conventional GWAS locus boundaries may also carry strong regulatory relevance in uterine tissue. Given that GWAS locus definitions are constrained by lead SNP selection and fixed window sizes, these findings suggest that outside-locus regulatory prioritization can complement standard locus-based analyses by identifying potentially functional variants not captured within established GWAS regions. The comparison of AlphaGenome-prioritized SNPs with multi-ancestry endometriosis GWAS SNPs [14] also supports the broader interpretation of regulatory modalities. Of the 37 multi-ancestry lead SNPs, 22 were already present in the 10,000 SNP GWAS dataset [3] and all belonged to the outside-locus category rather than to the 42 GWAS lead SNP windows. This suggests that SNPs located outside the original locus framework may subsequently be identified as endometriosis-associated signals as larger and more comprehensive GWAS datasets become available. Moreover, while in the multi-ancestry GWAS study, these SNPs were mapped to specific genes, AlphaGenome predicted multiple regulatory target genes at the same loci, including the previously mapped genes, thereby broadening the functional interpretation of these regions. Among these multi-ancestry SNPs, rs1799293, previously mapped to *ARHGAP26* gene was classified as tier 1 with support across six regulatory modalities. AlphaGenome additionally identified *FGF1* as regulatory target gene for this SNP alongside the previously mapped gene *ARHGAP26*. Promoter polymorphism-1385A/G (rs30411) at *FGF1* has been associated with endometriosis risk, with the A allele showing reduced frequency in endometriosis cases, implicating altered angiogenic signaling [61]. In parallel, *ARHGAP26* is downregulated in ectopic and eutopic endometrium compared with normal endometrium and was negatively associated with the severity of menorrhagia [62]. Together, the outside-locus and multi-ancestry GWAS comparison results suggest that functional interpretation may benefit from extending beyond previously defined lead-SNP windows. While the 42 published loci remain the central framework of this study, the outside-locus findings highlight additional genome-wide significant variants with predicted uterine regulatory effects. These variants may provide useful candidates for future fine-mapping, replication, and functional validation as larger GWAS datasets continue to refine the genetic architecture of endometriosis.

## Conclusions

This study demonstrates that integrating GWAS signals with uterus-specific regulatory predictions from AlphaGenome can refine the functional interpretation of endometriosis risk loci. By extending analysis beyond published lead SNPs to include additional GWAS-ranked and surrounding variants, we show that non-lead variants can exhibit stronger predicted regulatory effects and multimodal support across loci. This re-prioritization framework, supported by tissue-specific filtering and multi-layer regulatory annotation, improves the identification of likely causal non-coding variants and provides a more nuanced view of the genetic architecture underlying endometriosis. However, the findings are based on computational predictions and require experimental validation. In addition, uterus-specific filtering may not capture all disease-relevant cell types, and the strict prioritization threshold may exclude variants with moderate functional effects. The fixed locus window and LD assumptions may also miss long-range or ancestry-specific regulatory mechanisms. Overall, this approach provides a refined and scalable framework that can be incorporated into standard GWAS analysis pipelines to enhance variant prioritization and functional interpretation, thereby supporting more systematic identification of candidate causal variants in endometriosis and related complex traits.

## Abbreviations

ATAC-seq: Assay for Transposase-Accessible Chromatin sequencing
CAGE: Cap Analysis of Gene Expression
DNase-seq: DNase sequencing
GTEx: Genotype-Tissue Expression
GWAS: Genome-wide association study
PRO-cap: Precision Run-On coupled with cap analysis
RNA-seq: RNA sequencing
SNP: Single-nucleotide polymorphism
TF ChIP-seq: Transcription factor chromatin immunoprecipitation sequencing
eQTL: Expression quantitative trait locus
LD: Linkage disequilibrium
3D: Three-dimensional

## Declarations

### Ethics approval and consent to participate

Not applicable.

### Consent for publication

Not applicable.

### Data Availability

All data analysed and presented in this study are included in this published article and its additional files. Additional global AlphaGenome raw prediction outputs generated from the analysis of the 10,000 SNPs and the analysis code are available from the corresponding author upon reasonable request.

### Competing interests

The authors declare that they have no competing interests.

### Funding

The present study was funded by: Horizon Europe grant (NESTOR, grant no. 101120075) of the European Commission; Novo Nordisk Foundation (grant no. NNF24OC0092384); Estonian Research Council (grants nos. PSG1082, PRG1076); Swedish Research Council (grant no. 2024-02530); Estonian Ministry of Education and Research Centres of Excellence grant TK214 name of CoE and the Sigrid Jusélius Foundation.

### Authors’ contributions

Sanu Bifal Maji conceived the study design, performed the AlphaGenome-based variant annotation and prioritization analyses, conducted data processing, generated figures and tables, interpreted the results, and drafted the manuscript. Alberto Sola-Leyva, Apostol Apostolov, Lucia Blanco-Rodriguez contributed to data interpretation, methodological refinement, and manuscript revision. Andres Salumets and Amruta D. S. Pathare supervised the study, contributed to conceptualization and interpretation of the results, and critically revised the manuscript. All authors read and approved the final manuscript.

## Data Availability

All data produced in the present work are contained in the manuscript

## Notes

### Competing Interest Statement

The authors have declared no competing interest.

### Author Declarations

This study used ONLY openly available human data that were originally located in the supplementary information of the following articles: 1. Rahmioglu N, Mortlock S, Ghiasi M, et al. The genetic basis of endometriosis and comorbidity with other pain and inflammatory conditions. Nature Genetics. 2023;55:423-436. https://doi.org/10.1038/s41588-023-01323-z 2. Koller D, He J, Lokhammer S, et al. Multi-ancestry genome-wide association and integrated multi-omics analyses of endometriosis and its clinical manifestations. Nature Genetics. 2026. https://doi.org/10.1038/s41588-026-02582-2.

