## Additional file 1 for "Decoding the regulatory genetic architecture of endometriosis using AlphaGenome"

Figure 1: **AlphaGenome workflow for prioritizing endometriosis-associated SNPs**

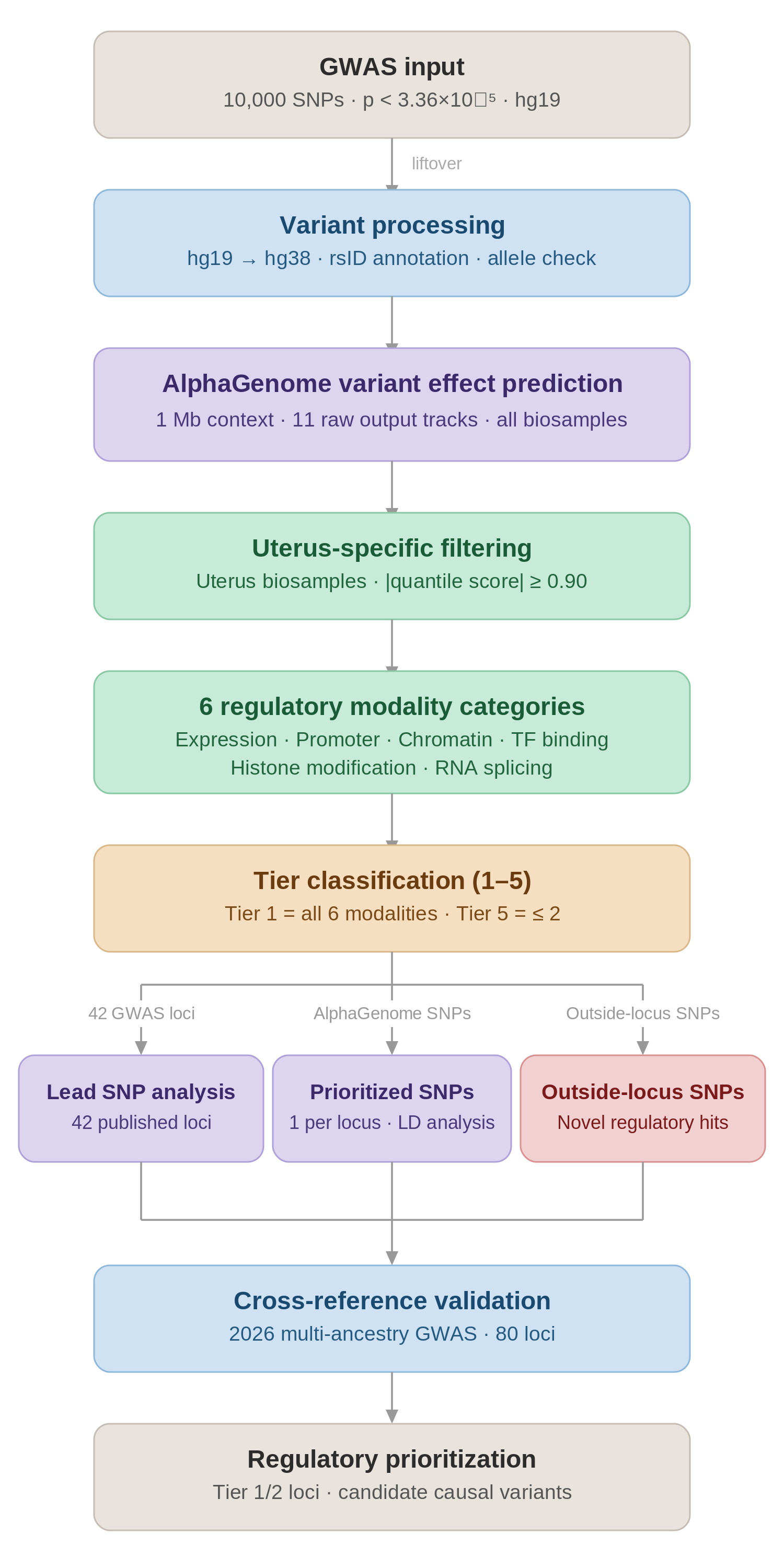

**Figure 1. AlphaGenome workflow for prioritizing endometriosis-associated SNPs**. A total of 10,000 endometriosis-associated GWAS SNPs (*p* < 3.36×10⁻⁵) were converted from hg19 to hg38 and analyzed using AlphaGenome with a 1 Mb sequence context across 11 regulatory output tracks. Predictions were restricted to uterus-specific biosamples, and high-confidence signals (|quantile score| ≥ 0.90) were retained. Regulatory outputs were grouped into six modalities gene expression, promoter activity, chromatin accessibility, transcription factor binding, histone modification, and RNA splicing. Each SNP was assigned a tier based on multimodal support (Tier 1 = all six modalities; Tier 5 = ≤2 modalities). Tiered SNPs were evaluated across three groups: published lead GWAS SNPs at 42 genome-wide significant loci, AlphaGenome-prioritized alternative SNPs within ±500 kb of these loci, and outside-locus SNPs. Results were further cross-referenced with a recent multi-ancestry endometriosis GWAS to validate novel regulatory signals.
